# Evaluating the Utility of UV Lamps to Mitigate the Spread of Pathogens in the ICU

**DOI:** 10.1101/2020.04.29.20085704

**Authors:** Andrew Gostine, David Gostine, Jack Short, Arjun Rustagi, Jennifer Cadnum, Curtis Donskey, Tim Angelotti

**Affiliations:** Northwestern Lake Forest Hospital, Lake Forest, IL; Cedar Sinai Medical Center, Los Angeles, CA; Stanford University Medical Center, Stanford, CA; Louis Stokes Cleveland VA Medical Center, Cleveland, OH

## Abstract

**Introduction:** Herd-immunity is the practice by which an immunized portion of a population or herd confers a measure of protection for individuals who have not been immunized. While commonly associated with vector vaccination, it offers the same practical approach to aseptic strategies in the hospital. In theory, the more decontaminated surfaces in a hospital, the greater immunity conferred to patients against the spread of nosocomial infections. The aim of this study was to evaluate the effectiveness of UV lights in preventing the spread of a DNA tracer from room-to-room in an ICU.

**Methods:** In a prospective trial, a non-pathogenic DNA virus was inoculated onto a variety of high-touch surfaces in an ICU patient room. Investigators swabbed frequently touched surfaces in non-inoculated ICU rooms at 24, 48, and 96-hours post inoculation. Culture specimens were analyzed for the presence of viral DNA via PCR. After baseline data was obtained, focused UV lights were deployed to cover frequently touched surfaces in the ICU including vitals monitors, ventilators, keyboards, dialysis machines and IV pumps. Inoculation and culturing were then repeated.

**Results:** Prior to UV implementation, the DNA tracer disseminated to 10% of tested surfaces in noninoculated rooms at 48 hours. Post UV light deployment, only 1.2% of surfaces tested positive for the DNA tracer after 48 hours.

**Conclusion:** UV decontamination significantly and meaningfully retarded the spread of the mosaic virus DNA across ICU surfaces with a relative reduction in 90% of surfaces contaminated at 48-hours from 10.10% of surfaces pre-UV to 1.20% of surfaces post-UV (p < 0.0001). Realtime UV decontamination holds the potential to confer protection to ICU patients by reducing the number of surfaces that can serve as a nidus for infection transmission.

**Highlights:**

- Even with standard cleaning, a mosaic virus spread to 10% of surfaces in the ICU.
- Post UV light deployment, only 1.2% of surfaces tested positive for the DNA tracer.
- UV light retarded the spread of DNA virus with a reduction of 90% at 48-hours.

## Background

The Center for Disease Control (CDC) estimates 1 in 31 hospitalized patients suffer a healthcare-associated infection (HAI) annually.^1^ The economic implications of HAIs are far reaching and cost the medical system between $35.7 and $45 billion each year.^2,3,4^ In one meta-analysis looking at additional hospital costs per-case, central line–associated bloodstream infections incurred an additional $45,814 (95% CI, $30,919-$65,245) to the total hospital stay, followed by ventilator-associated pneumonias at $40,144 (95% CI, $36,286-$44,220), surgical site infections at $20,785 (95% CI, $18,902-$22,667), Clostridium difficile infections at $11,285 (95% CI, $9,118-$13,574), and catheter-associated urinary tract infections at $896 (95% CI, $603-$1,189).^5^ A commonality among many HAIs is their association with these devices, e.g., lines, catheters, and ventilators.^6^

Portable equipment and other shared devices, e.g., keyboards, touchscreens, and pens, may be an underappreciated source of transmission of healthcare-associated pathogens; these items are often contaminated by microbes and cleaning may be suboptimal.^7^ In several outbreak investigations, shared equipment has been implicated as a fomite for transmission of pathogens.^8^ One study from a laboratory at the University of Siena analyzed keyboards in a shared working space and found microbes in counts ranging from 6 CFU/key to 430 CFU/key, including staphylococcus, streptococcus, and enterococcus species.^9^ Thus, it is plausible that decreasing the rate of cross-contamination of hospital equipment might be an effective strategy to mitigate the spread of HAIs.^10,11,12^ This idea can be approached in the conceptual framework of herd-immunity, that is, the practice by which an immunized portion of a population confers a measure of protection for individuals who have not been immunized.^13^ While commonly associated with vaccines, this paper explores the application of herd immunity in aseptic strategies within the hospital where reducing the prevalence of contaminated surfaces should confer protection against pathogen transmission.

Since first described in 1903 by Nobelist Niels Finsen, ultra-violet (UV) light has been recognized as an effective decontamination technique.^14,15^ UV light works by disrupting the structure of the microorganism’s DNA^16,17^ and is a well-validated sterilization tool. In one study looking at the role of UV light in stethoscope sterilization, UV-C demonstrated the capacity to maintain high levels of disinfection against common HAI microorganisms.^18^ Historically, the deployment of UV light in the presence of humans required significant measures to protect staff and patients from exposure. Recent developments in UV technology has yielded a new class of devices, capable of monitoring for user input and delivering low dose of UV-C light when human exposure is not detected. Each dose is set to a safe and optimal disinfection cycle to eliminate pathogens with minimal human exposure.^17^ Should a human need to interact with the device being sterilized, the UV light automatically pauses its operation, waits for use to cease, and then resumes the cleaning cycle.

A 2016 study in the American Journal of Infection Control demonstrated the utility of low-intensity UV-C devices in reducing bioburden on hospital computer keyboards.^19^ In the current study, we investigated the capacity of these UV-C devices in reducing the spread of a mosaic virus DNA tracer, i.e., a proxy for pathogens, across hospital rooms in the ICU.

## Materials and Methods

### Study Site and Design

The trial was carried out via a pre-post experimental design. The study site was a single, 33-bed mixed medical and surgical intensive care unit at an academic medical center in Stanford, CA. Study approval was obtained from Stanford University Human Subjects Panel (IRB Protocol #45006). Faculty and staff were educated on the function of the UV lights prior to implementation, however, there was no instruction to alter staff behavior or decontamination practices within the ICU.

### UV Light Installation and UV Treatment Protocol

The manufacturer (UV Partners, Inc., Grand Haven, MI USA) installed the UV lights (UV Angel) on 140 high-touch devices in the ICU. These devices included: IV pumps (Alaris Pump, BD Medical, Franklin Lakes, NJ USA), stationary computer keyboards, portable computer workstation surfaces and keyboards, touchscreen vitals monitors, ventilators, and Pyxis drug dispensers (Pyxis Medstation, BD Medical, Franklin Lakes, NJ USA). The UV lights used were small, 3.2 cm deep by 30.5 cm wide, and were placed above high-touch items with the goal of providing fully automated decontamination cycles after each use. Detailed descriptions of the UV device have been published previously.^20^ The UV lights were programmed to turn on 90-seconds after user input was no longer detected. The UV light remained on for an 18-minute cycle. The cycle length was determined based on a previous analysis in which UV-C light was used to inactivate *C. diff* spores^21^. The UV Angel team collected data on the number of UV light cleaning cycles but otherwise did not intervene in the study proceedings. No other HAI reduction strategy, outside of the standard of care, were implemented during this analysis.

### Mosaic Virus Transmission

Pathogen transmission was performed using two genetically distinct mosaic virus DNA markers. Prior to installation of the UV lights, a standard quantity of cauliflower mosaic virus DNA was suspended in sterile saline. A standardized 1.15mL volume was sprayed onto high touch surfaces, including bed rails, touchscreen monitors, computer keyboards, and ventilators in 4 of the 33 ICU patient bays. Swabs from high-touch surfaces in the 29 non-inoculated bays were then obtained 24-hours (n = 100 swabs), 48-hours (n = 99 swabs) and 96-hours (n = 78 swabs) post inoculation to demonstrate the baseline spread of this pathogen proxy throughout the ICU. A total of 277 baseline swabs, plus positive controls, were obtained and amplified via polymerase chain reaction (PCR). After deployment of the UV lights, the same protocol was followed with a second, genetically distinct, mosaic virus DNA marker to determine the impact on pathogen transmission to non-inoculated bays. A total of 261 swabs plus positive controls were collected post-intervention at 24-hours (n = 98 swabs), 48-hours (n = 83 swabs) and 96-hours (n = 80 swabs).

### Viral Swabs and Processing

Viral swab collection throughout the study was performed by a trained operator using Copan eSwab collection kits (Copan Diagnostics, Inc., Murrieta, CA USA). The swab tip was dipped in the kit’s modified liquid transport medium inside the vial, and the remainder of the liquid was discarded to prevent dilution of the mosaic virus. The swab was passed multiple times in a wide pattern over the collection surface and inserted back into the collection tube. The applicator tip was broken off into the vial and the screw cap sealed. The specimen was then immediately labeled according to protocol. Researchers and lab staff were not blinded to the surface or whether or not the samples came from UV protected devices.

Throughout both phases of the study, the swab processing was consistent in methodology. The same investigator conducted all portions of the collections. Storage and shipping, which consisted of insulated boxes with ice packs, were consistent for all swabs and expedited to the research lab for analysis at the earliest possible time. The swab collection technique, labeling, and volume of liquid culture medium used to gather samples were consistent throughout all phases.

### Device Touch and Cycle Analysis

During the 96-hour deployment of the UV lights on various high touch surfaces, motion sensors in the UV lights recorded the number of unique touches on UV-protected devices. Based on a prior study from 2016, we defined a unique touch as occurring at least 90 seconds after the previous interaction completed.^19^ This length of time was chosen based on the plot of the percent of touches occurring versus length of time since the last touch. It revealed a logarithmic curve with slightly over 50% of touches occurring in the first 90 seconds. Beyond that point, the curve became approximately linear suggesting a uniform probability of device use after 90 seconds. The results of the protected device touches per 24-hour period are displayed in Table 1 with an average of 64 touches per device class per 24-hour period. Mobile workstation surfaces and keyboards were touched the most followed by Pyxis machines and IV pumps. Ventilators and vitals monitors were touched the least.

**Table 1:**
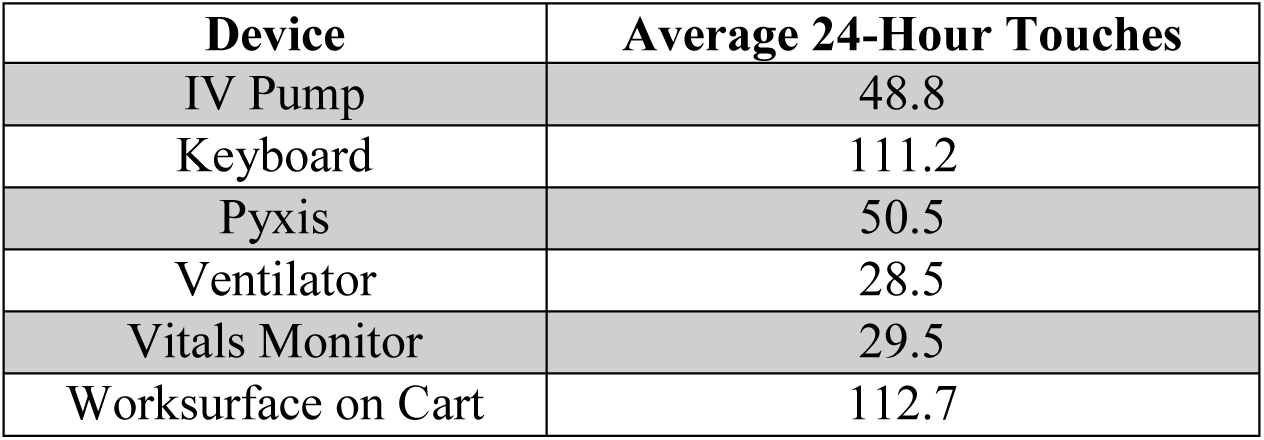
Estimated Number of Unique Device Touches per 24-Hour Period

### Statistical Analysis

We used descriptive statistics for comparison, in charts and in graphs, to analyze viral transmission interruption. The Wilcoxon signed-rank two-tailed test was used to compare the pre and post-intervention results. Analysis was done using XLSTAT Version 2019.3.2 (Copyright Addinsoft (2019) XLSTAT and Addinsoft are Registered Trademarks of Addinsoft)

## Results

### Mosaic Virus Transmission

When analyzing the data of the mosaic virus dissemination in the absence of UV-C treatment, the virus disseminated to 2.00% (n = 2 of 100) of surfaces after 24 hours, peaked at 10.10% (n = 10 of 99) after 48 hours, and was found on 5.13% (n = 4 of 78) of tested surfaces after 96 hours (Figure 1). Post-UV deployment, 0.00% of surfaces tested positive for the mosaic virus DNA after 24 hours, 1.20% (n = 1 of 83) of sites tested positive for the mosaic virus DNA after 48 hours, and 2.50% (n = 2 of 80) tested positive after 96 hours (Figure 1)

**Figure 1:**
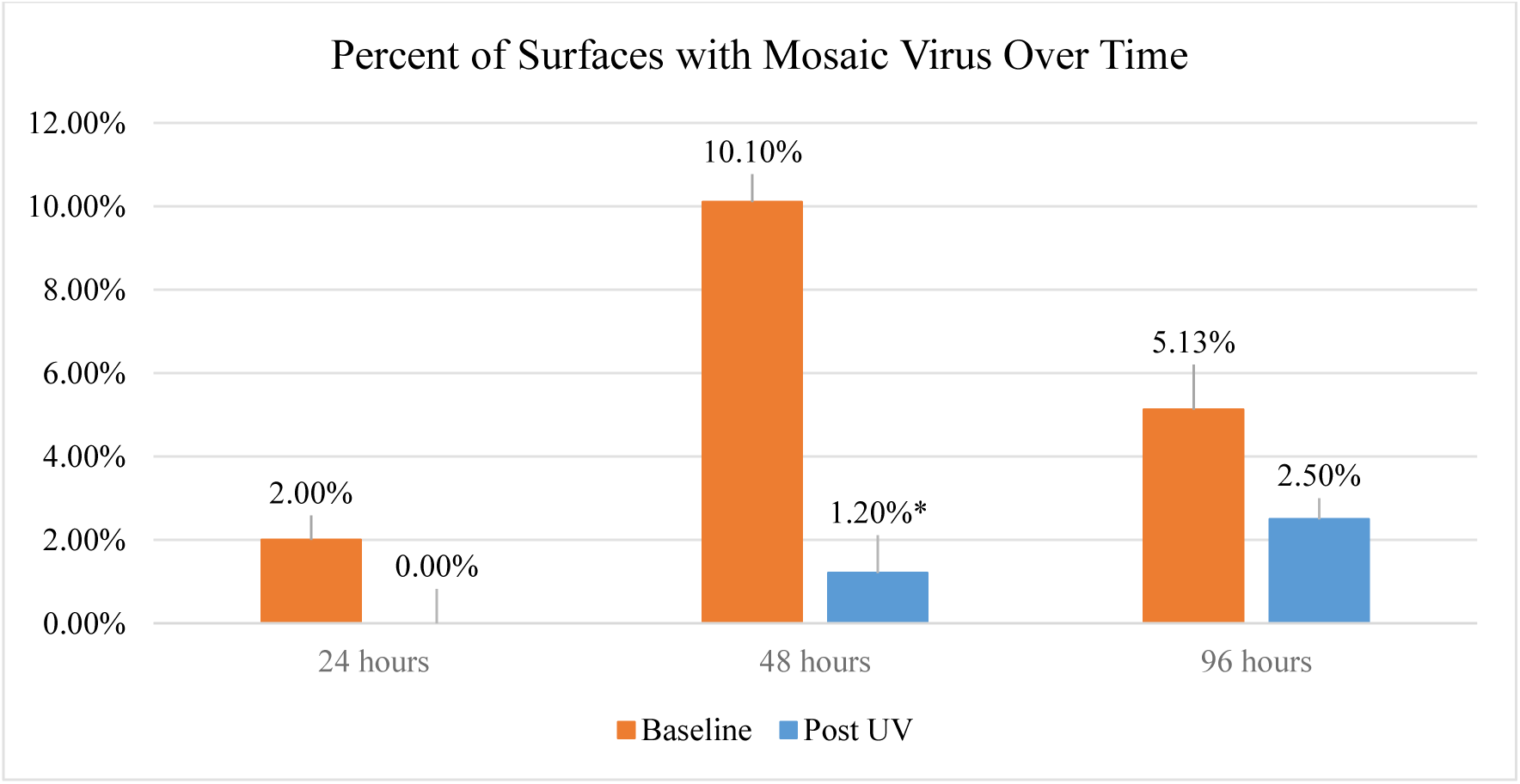
Percent of ICU Surfaces Contaminated with Mosaic Virus at Sequential Samplings *\*Denotes statistically significant reduction*

The greatest reduction in mosaic virus transmission post-UV implementation was observed at 48 hours with a 90% relative reduction (p < 0.0001) in mosaic virus spread across surfaces in the ICU (Table 2).

**Table 2:** Percent of Surfaces Bays Contaminated with Mosaic Virus at Sequential Samplings

| Hours post Inoculation | 24 | 48 | 96 |
| --- | --- | --- | --- |
| Pre-UV Surfaces Positive (Percent) | 2.00% | 10.10% | 5.13% |
| Pre-UV Surfaces Positive (Number) | N = 2 | N = 10 | N = 4 |
| Post UV Surfaces Positive (Percent) | 0% | 1.20% | 2.50% |
| Post-UV Surfaces Positive (Number) | N = 0 | N = 1 | N = 2 |
| Significance | p = 0.125 | p < 0.0001 | p = 0.688 |

## Discussion

HAIs are commonly spread through contaminated hospital surfaces, air, water, or providers that serve to transport pathogens.^22,23^ Countless surfaces in hospitals are reservoirs for viable pathogens including MRSA and VRE. In rooms of patients with diarrhea, viable MRSA has been collected from 59% of the room surfaces and viable VRE has been collected from 46% of room surfaces.^24^

Despite the known prevalence of contaminated surfaces, current cleaning practices are inadequate.^25^ Terminal room cleaning after patient discharge only reduced bacterial contamination to undetectable levels on 49% of surfaces, including less than 30% for toilet hand holds, bedpan cleaners, room doorknobs, and bathroom light switches.^26^ Furthermore, these cleaning practices are, at best, occurring once per day.^27^ These high-touch surfaces serve as potent sources for bacterial and viral transmission and are concerning given the ratio of touches to cleanings that they receive.^28^ In one analysis, it was noted that while nearly two-thirds of clinical staff touched surfaces within a patient’s room these same surfaces were only cleaned by environmental services a maximum of once per day.^29^ Alarmingly, it is these same surfaces nearest to the patient that confer the highest infection risk.^30,31,32^

Despite numerous studies demonstrating its utility, UV light use remains underutilized in the hospital, in part due to issues regarding costs, safety, or staffing needs.^33^ The small UV devices used in the current study have some advantages and disadvantages over larger UV room decontamination devices. The devices are intended to be used when people are present and are fully automated. In a previous study, we found that the UV devices effectively decontaminated keyboards with no interruption of workflow, no additional staffing, and no adverse effects due to UV exposure. This follow-up study again demonstrated the capacity of UV light to reduce bacterial burden on some of the most commonly touched surfaces in the hospital.

It seems reasonable to conclude that UV treatment could help mitigate the spread of infectious agents by reducing cross-contamination from commonly used objects in the ICU, given the frequency with which these devices are used. We approached this idea using the conceptual framework of herd immunity, in which immunization of the majority confers immunity to the minority. While most often associated with vaccination of humans, the transmission modeling can be extended to decontamination practices in the hospital. Here, we theorized that every physical contact between a staff member and a high-touch surface was an interaction in need of vaccination, i.e., via the application of passive UVC light. By providing real-time pathogen reduction technology to various fomites, we reduced the probability of a mosaic virus to spread within the ICU. In combination with high handwashing rates, which were assumed to be constant throughout the study, as the number of decontaminated surfaces in the hospital increases, the number of viable transmission routes between patients subsequently decreases.^34^ Greater utilization of validated aseptic techniques like UV decontamination could enhance patient safety and improve outcomes, especially in critical ill and otherwise susceptible patients.^35^

This study has limitations given the resources and time available to run the trial. While we significantly reduced the transmission of pathogens on the UV protected devices, we do not know what percent of interactions those high-touch devices represent as it is impractical to count every touch that occurs in the ICU. Given this limitation, we only tested known high-touch surfaces. These surfaces were identified by a review of time in motion studies.^36^ We also did not completely eliminate the spread of the mosaic virus and are, therefore, unable to calculate a fomite “vaccination threshold” for the complete protection of ICU patients. Lastly, we were not able to consent patients for culturing, so it was not within the means of this analysis to provide direct evidence that the mosaic virus was transmitted to patients from the numerous in room surfaces. We were only able to show definitively that the mosaic virus moved from one surface to the next. Further study is needed to examine if this causal reduction in pathogen transmission leads to a reduction in specific healthcare-associated infections.

## Conclusion

This study confirmed our hypothesis. UV decontamination significantly and meaningfully retarded the spread of the mosaic virus across ICU surfaces with a relative reduction of 90% at 48-hours from 10.10% of tested surfaces to 1.20% of surfaces post-UV (p < 0.0001). Realtime UV decontamination holds the potential to confer protection to ICU patients by reducing the number of surfaces that can serve as a nidus for infection transmission.

## Data Availability

Raw data available upon request.

